# Estimated Incidence of Respiratory Syncytial Virus (RSV)-related Hospitalizations for Acute Respiratory Infections (ARIs), including Community Acquired Pneumonia (CAP), in Adults in Germany

**DOI:** 10.1101/2024.06.09.24308507

**Authors:** Caihua Liang, Elizabeth Begier, Stefan Hagel, Juliane Ankert, Liz Wang, Claudia Schwarz, Lea J. Bayer, Christof von Eiff, Qing Liu, Jo Southern, Jeffrey Vietri, Sonal Uppal, Bradford D. Gessner, Christian Theilacker, Mathias Pletz

## Abstract

**Background:** RSV is a leading cause of ARI, including CAP, in older adults, but available data often substantially underestimate incidence. We estimated RSV-related ARI hospitalization incidence from a prospective CAP study and adjusted for undiagnosed RSV infections due to use of nasopharyngeal/nasal swab testing only.

**Methods:** We conducted active, population-based surveillance of adult CAP hospitalizations in Thuringia (Germany) between 2021–2023. Participant nasopharyngeal/nasal swabs were RSV-tested by multiplex nucleic acid amplification testing. To estimate RSV-related CAP incidence, age-group specific proportions of RSV positivity among tested patients were applied to all-cause CAP incidence. To adjust for underdiagnosis due to nasopharyngeal/nasal swab sampling only and the percentage of ARI with pneumonia diagnoses, we used data from a large, prospective, multispecimen study assessing impact of collecting multiple specimens (nasopharyngeal/nasal swab, saliva, paired serology, and sputum) among 3,669 adults hospitalized for ARI.

**Results:** Among 1,040 enrolled adults (≥18 years) with radiologically confirmed CAP, 38 tested RSV-positive via nasopharyngeal/nasal swab (3.7%). The percentage positive increased to 7.8% after adjusting for higher RSV detection with multiple specimens compared to nasopharyngeal/nasal swab only. Adjusted RSV-related CAP hospitalization rates were 4.7 (95%CI 1.5–11.2) and 109.1 (95%CI 89.6–131.6) per 100,000 adults aged 18–59 and ≥60 years, respectively. Adjusted incidences of RSV-related ARI were 18.4 (95%CI 11.0–28.9) and 377.6 (95%CI 340.5–417.7) per 100,000 adults aged 18–59 and ≥60 years, respectively. Among RSV-positive CAP hospitalizations, 12.1% of patients aged ≥65 years died within 30 days, with no deaths in those aged 18–64 years. Cardiovascular events occurred in 11.1% of patients aged 18–64 and 36.4% of those aged ≥65 years.

**Conclusions:** Older adults in Germany face a high burden of RSV-related ARI hospitalizations, including CAP, underscoring RSV vaccination’s potential utility for this population.

**KEY PUBLIC HEALTH MESSAGE:** *What did you want to address in this study and why?:* Hospital administrative data significantly underestimate respiratory syncytial virus (RSV) incidence due to infrequent testing and lower sensitivity of single nasopharyngeal/nasal swab testing among adults. No prospective incidence studies are available for Germany and most other European countries. We aimed to estimate RSV-related acute respiratory infection (ARI) hospitalization incidence from a prospective community-acquired pneumonia (CAP) study and adjust for undiagnosed RSV infections due to limited testing and use of nasopharyngeal/nasal swab testing only. Detailed data on RSV disease burden are crucial for developing vaccination policies.

*What have we learnt from this study?:* Adjusted annual incidence of RSV-related ARI rates were 18.4 (95% CI 11.0–28.9) and 377.6 (95% CI 340.5–417.7) per 100,000 population for adults 18–59 and ≥60 years, respectively. Among RSV-positive CAP hospitalizations, 12.1% of patients aged ≥65 years died within 30 days, with no deaths in those aged 18–64 years. Cardiovascular events occurred in 11.1% of patients aged 18–64 and 36.4% of those aged ≥65 years.

*What are the implications of your findings for public health?:* Our findings are similar to recent time-series incidence results from Germany (236–363/100,000 for adults ≥60 years) and underscore the substantial burden of RSV among adults, particularly the high rate of cardiovascular events contributes to a probably underestimated burden of RSV disease.

## Introduction

Lower respiratory tract infections (LRTIs) are leading causes of morbidity and mortality globally [1, 2]. Pneumonia is the most common diagnosis of hospitalized acute lower respiratory tract disease (aLRTD), accounting for 31% of aLRTD in a UK study [3] and 29% in a US prospective cohort study [4]. In Germany, a recent retrospective cohort study using a representative healthcare claims database reported an incidence of hospitalized community-acquired pneumonia (CAP) 1,061 per 100,000 person-years in adults aged ≥60 years [5].

Respiratory syncytial virus (RSV) is now being recognized as one of the leading causes of acute respiratory infections (ARIs) in older adults, including among patients with pneumonia [6, 7]. A recent global systematic literature review and metanalysis reported a pooled adjusted RSV-related ARI hospitalization incidence of 347 per 100,000 for adults aged ≥65 years, corresponding to 787,000 annual hospitalizations among older adults in high-income countries [8]. In the US, roughly 2–9% of older adult pneumonia hospitalizations have been attributed to RSV [6, 9] and this fraction increases to up to 15% during the winter months depending on the diagnostic approaches used and the season under investigation [4, 10, 11]. RSV-related ARI hospitalizations include pneumonia, but also non-pneumonic LRTI and exacerbations of cardiopulmonary disease caused by RSV infection. In one study in Finland, pneumonia accounted for 14% (3/22) of RSV-related ARI hospitalizations [4]. A more recent prospective study using multiple specimen types found pneumonia to comprise 28% (71/254) of RSV-related ARI hospitalizations when multiple additional specimen types (i.e., sputum, saliva and blood) were added rather than relying on nasopharyngeal (NP)/nasal swab alone [12].

Accurate estimation of RSV-related hospitalization incidence is challenging due to several causes of RSV case underascertainment [13]. RSV clinical presentation is non-specific and thus diagnostic testing is necessary for confirmation of infection [10]; however this is infrequently performed because it usually does not impact clinical management [13, 14]. When performed, RSV laboratory testing may miss 50% of infections if only a single specimen is used for diagnostic testing (often NP/nasal swab reverse transcription polymerase chain reaction [RT-PCR]) [15, 16]. This is likely due to viral titers in respiratory secretions in adults being typically lower than titers in children, so that RSV may no longer be detectable at the time of testing at presentation/hospital admission [17], which tends to occur later in illness than for other respiratory viruses [13].

Current assessments of RSV burden focus on RSV-related ARI hospitalizations, conducted either through prospective studies utilizing direct diagnostic testing, retrospective studies using diagnosis codes from large administrative databases, or through time-series modeling approaches. Due to the reduced sensitivity of a single RT-PCR NP/nasal swab, prospective estimates should be adjusted by the percentage increase in RSV detections associated with using multiple diagnostic specimen types [8, 18]. Estimates based on diagnosis/ICD codes only have been shown to substantially underestimate the RSV incidence globally and in Germany [18-20]. Model-based studies link the variations in a broad definition of respiratory disease (e.g. all respiratory diseases) to the variation in RSV activity to estimate the proportion of respiratory disease attributable to RSV. This non-specific outcome definition can account for the undiagnosed RSV cases, which might have been coded as other diseases [20, 21]. However, results can vary by different modeling approaches and viral activity proxies selected and prospective incidence studies are considered the gold standard.

Currently, no RSV-related hospitalization estimates based on active surveillance and prospective testing are available for Germany. We used data from a prospective study of CAP incidence [22] to estimate RSV-related CAP and ARI hospitalization incidence, including adjustment for undiagnosed RSV infections due to use of NP/nasal swab testing only and non-CAP ARI events.

## Methods

### Study design and study population

The CAP incidence study is a population-based multi-hospital, prospective study to determine the CAP hospitalization incidence rate among adults aged ≥18 years in Thuringia, Germany. Active surveillance for CAP has been conducted at three hospitals: Jena University Hospital (JUH); the SRH Wald-Klinikum Gera; and the SRH Zentralklinikum Suhl. Study period for this analysis is July 2021–June 2023 (24 months), encompassing two winter seasons. Adults who met the following inclusion criteria were offered enrollment: aged ≥18 years at the time of hospital admission with CAP and had evidence of pneumonia within first 48 hours of hospital admission based on radiologic findings consistent with pneumonia and presence of ≥2 of following 10 clinical signs or symptoms (fever within 24 hours before enrollment; hypothermia within 24 hours of enrollment; chills or rigors; pleuritic chest pain; new or worsening cough; sputum production; dyspnea; tachypnea; malaise; abnormal auscultatory findings suggestive of pneumonia). Patients with radiologically-confirmed CAP were included in the analysis. The number of unenrolled persons meeting inclusion/exclusion criteria was recorded to allow for enumeration of all CAP hospitalizations regardless of enrollment status.

### Data sources

NP/nasal swabs (or alternatively mid-turbinate nasal swabs) were collected from enrolled participants and tested by multiplex nuclear acid amplification test (NAAT) for the presence of respiratory viruses, including RSV.

The following data were collected from the study participants and hospital records on admission: age, gender, study period, smoking status, alcohol use and chronic medical conditions. The risk level was classified as low risk, at-risk and high-risk using CDC definitions for pneumococcal disease [23], which are similar to risk factors for severe RSV infection [24, 25]. Cardiovascular events were defined as any of the following: new or worsening heart failure, new or worsening arrhythmia, acute coronary syndrome, cerebrovascular accident, deep venous thrombosis or pulmonary embolism, or cardiovascular-related death.

### Definition of Catchment Area Population

To estimate the RSV-related CAP incidence, the catchment area population was defined based on residential zip codes. As uninterrupted CAP surveillance with full case capture was only available from JUH, incidence estimations are based on this study site only. The total population of adults aged ≥18 years from catchment area zip codes formed the denominator of the incidence calculation. The observed unadjusted incidence rates were multiplied by the inverse of the hospital market share in the catchment area (i.e., 97%) to adjust for healthcare seeking at other facilities by residents of the catchment area.

### Statistical analysis

Demographics and risk status were descriptively summarized for all patients meeting the case-definition for CAP, according to testing status (tested and untested) and regardless of their enrollment status. All variables were classified as either binary or categorical and are presented as counts with percentages. The RSV positive proportion from NP/nasal swab alone (unadjusted) was calculated as the number of RSV positive cases divided by the total number of participants who had a valid NP/nasal swab test result. In order to adjust for underascertainment of RSV detection based on use of NP/nasal swab testing alone, we used results from the previously published prospective study detecting RSV using multispecimen types (referred to as multispecimen study hereafter) [12]. The percent increase in RSV detection rate associated with additional specimens was calculated using the positive proportion from NP/nasal swab plus other specimens (i.e., saliva, paired serology, and sputum) and the positive proportion from NP/nasal swab alone. This percent increase among different age groups served to correct for underascertainment of RSV cases due to missed RSV infections associated with nasopharyngeal/nasal swab diagnostic testing only.

The unadjusted RSV-related CAP incidence rates were calculated by multiplying age-group specific proportions of RSV detection among tested patients with all-cause CAP incidence for all eligible participants enrolled at Jena site, based on the assumption that RSV detection rate would not differ significantly between those tested and untested. The adjusted RSV-related CAP incidence rates were calculated using the percent increase in RSV detection obtained from the multi-specimen study [12]. This was achieved by multiplying percent increase associated with using multiple diagnostic testing specimens with the unadjusted RSV-related CAP incidence.

Incidence rate of RSV-related ARI hospitalizations (both unadjusted and adjusted) was then estimated by dividing the RSV-related CAP incidence rates (unadjusted and adjusted) by the proportions of CAP among RSV-associated ARI hospitalizations by age group, obtained from the multi-specimen study [12]. The corresponding 95% confidence intervals (CIs) were calculated by the exact method using Poisson distribution. R version 4.3.0 and SAS version 9.4 (SAS 9.4, Cary, NC. SAS Institute Inc.) were used to perform all analyses.

### Ethics/Ethical Approval

All study participants or legal representatives were informed about the study and consented to the participation. Local ethics committees approved the conduct of the study (2020-1947-material).

## Results

At JUH, there were 293 patients hospitalized with CAP tested for RSV by NP/nasal swab and 741 were not tested (including enrolled and non-enrolled patients). Patients not tested were slightly older (84.2% aged ≥65 years vs 77.5%), had lower prevalence of high-risk conditions (38.7% vs 58.4%) but a higher prevalence of at-risk conditions (52.8% vs 35.5%) than tested individuals (Table 1).

**Table 1.** Characteristics of adults hospitalized at Jena University Hospital with radiologically-confirmed CAP patients by RSV testing status.

|  | NP/Nasal Swab Tested |  | NP/Nasal Swab Untested*** |  | P-Value |
| --- | --- | --- | --- | --- | --- |
|  | N | % | N | % |  |
| **Overall** | 293 | 100 | 741 | 100 |  |
| **Age group** |  |  |  |  |  |
| 18-59 years | 38 | 13 | 73 | 9.9 | 0.145 |
| ≥60 years | 255 | 87 | 668 | 90.1 | 0.145 |
| 18-64 years | 66 | 22.5 | 117 | 15.8 | 0.010 |
| ≥65 years | 227 | 77.5 | 624 | 84.2 | 0.010 |
| **Sex** |  |  |  |  |  |
| Female | 125 | 42.7 | 309 | 41.7 | 0.778 |
| Male | 168 | 57.3 | 432 | 58.3 |  |
| **Study period\*** |  |  |  |  |  |
| Year 1 | 135 | 46.1 | 341 | 46 | 0.987 |
| Year 2 | 158 | 53.9 | 400 | 54 | 0.987 |
| Overall study period | 293 | 100 | 741 | 100 |  |
| **Risk status\*\*** |  |  |  |  |  |
| Low-risk | 18 | 6.1 | 62 | 8.4 | 0.228 |
| At-risk | 104 | 35.5 | 391 | 52.8 | <0.001 |
| High-risk | 171 | 58.4 | 287 | 38.7 | <0.001 |

Abbreviations: CAP=community-acquired pneumonia; CDC = centers for disease. NP/nasal swab=nasopharyngeal (NP)/nasal swab
\* Year 1: 01Jul2021 – 30Jun2022; Year 2: 01Jul2022 – 30Jun2023
\*\* The risk status was classified using CDC definitions for pneumococcal disease [23]
\*\*\* include enrolled and non-enrolled patients

### RSV detection among CAP patients

Among 1,040 radiologically confirmed CAP cases enrolled and tested at the three study sites between 01 July 2021 and 30 June 2023, 38 CAP cases had RSV detected, with an overall positive proportion of 3.7% among adults aged ≥18 years (Supplementary Table S1). RSV positive proportions were higher in the older age group (4.1% vs 2.4% for adults aged ≥60 years and 18–59 years, respectively) and were generally higher in Year 2 (01. July 2022–30. June 2023) (5.5%) than Year 1 (01. July 2021–30. June 2022) (2.3%), however, they were similar across risk status (3.3% for low-risk, 3.5% for at-risk and 3.9% for high-risk). After adjustment for underascertainment of RSV cases, the overall positive proportion increased from 3.7% to 7.8%, corresponding to a 112% increase in RSV detection resulting from adding multiple specimen results (i.e., saliva, paired serology, and sputum) beyond NP/nasal swab (Table 2).

**Table 2.** Unadjusted and adjusted incidence rates of RSV-related CAP and ARI hospitalizations in Thuringia/Germany 2021–2023.

| Age group (Years) | CAP incidence rate (per 100, 000) <sup>1</sup> | RSV positive proportion based on NP/nasal swab <sup>1</sup> | Percentage Increase in RSV detection <sup>2</sup> | RSV-related CAP incidence rate per 100,000 (95% confidence interval) <sup>3</sup> |  | Proportion of CAP in RSV-related ARI hospitalization <sup>5</sup> | RSV-related ARI incidence rate per 100,000 (95% confidence interval) <sup>6</sup> |  |
| --- | --- | --- | --- | --- | --- | --- | --- | --- |
|  |  |  |  | Unadjusted | Adjusted <sup>4</sup> |  | Unadjusted | Adjusted |
| Year 1: 01Jul2021 – 30Jun2022 |  |  |  |  |  |  |  |  |
| ≥18 | 451.0 (410.0, 494.5) | 2.3% | 112% | 10.4 (5.1,18.9) | 22.0 (13.8, 33.3) | 28.0% | 37.0 (26.1, 51.0) | 78.5 (62.1, 97.9) |
| 18-59 | 87.7 (70.6, 108.4) | 2.3% | 148% | 2.0 (0.2,7.2) | 5.0 (1.6, 11.7) | 25.4% | 7.9 (3.4, 15.6) | 19.7 (12.0, 30.5) |
| ≥60 | 1188.0 (1121.8, 1257.1) | 2.3% | 101% | 27.3 (18.0,39.6) | 54.9 (41.3, 71.5) | 28.9% | 94.5 (76.4, 115.6) | 190.0 (163.9, 219.0) |
| 18-64 | 119.4 (98.6, 142.4) | 1.7% | 141% | 2.0 (0.2, 7.2) | 4.9 (1.6, 11.5) | 22.6% | 9.0 (4.1, 17.1) | 21.6 (13.5, 32.8) |
| ≥65 | 1384.5 (1312.5, 1458.3) | 2.7% | 95% | 37.4 (26.4, 51.5) | 72.9 (57.1, 91.7) | 31.8% | 117.6 (97.3, 140.9) | 229.2 (200.5, 260.9) |
| Year 2: 01Jul2022 – 30Jun2023 |  |  |  |  |  |  |  |  |
| ≥18 | 528.7 (485.0, 575.9) | 5.5% | 112% | 29.1 (19.5, 41.8) | 61.6 (47.2, 79.0) | 28.0% | 103.9 (84.9, 125.9) | 220.2 (192.1, 251.3) |
| 18-59 | 69.3 (53.7, 87.3) | 2.6% | 148% | 1.8 (0.2, 6.9) | 4.5 (1.4, 11.0) | 25.4% | 7.1 (2.9, 14.6) | 17.6 (10.4, 28.0) |
| ≥60 | 1460.6 (1387.5, 1537.3) | 6.1% | 101% | 89.1 (71.6, 109.6) | 179.1 (153.8, 207.3) | 28.9% | 308.3 (274.8, 344.7) | 619.7 (571.9, 670.5) |
| 18-64 | 115.6 (95.9, 139.1) | 4.2% | 141% | 4.9 (1.6, 11.5) | 11.7 (6.0, 20.6) | 22.6% | 21.5 (13.4, 32.7) | 51.8 (38.7, 68.0) |
| ≥65 | 1691.7 (1613.0, 1773.9) | 6.0% | 95% | 101.5 (82.7, 123.3) | 197.9 (171.3, 227.5) | 31.8% | 319.2 (285.1, 356.2) | 622.4 (574.5, 673.3) |
| Overall study period: 01Jul2021 – 30Jun2023 |  |  |  |  |  |  |  |  |
| ≥18 | 489.8 (460.4, 520.6) | 3.7% | 112% | 18.1 (10.7, 28.6) | 38.4 (27.2, 52.6) | 28.0% | 64.7 (49.9, 82.5) | 137.2 (115.2, 162.2) |
| 18-59 | 78.5 (64.6, 94.5) | 2.4% | 148% | 1.9 (0.2, 7.1) | 4.7 (1.5, 11.2) | 25.4% | 7.4 (3.1, 15.0) | 18.4 (11.0, 28.9) |
| ≥60 | 1324.3 (1241.3, 1411.3) | 4.1% | 101% | 54.3 (40.8, 70.8) | 109.1 (89.6, 131.6) | 28.9% | 187.9 (162.0, 216.8) | 377.6 (340.5, 417.7) |
| 18-64 | 117.5 (101.1, 135.8) | 2.5% | 141% | 2.9 (0.6, 8.6) | 7.1 (2.9, 14.6) | 22.6% | 13.0 (6.9, 22.2) | 31.3 (21.3, 44.4) |
| ≥65 | 1538.1 (1437.9, 1643.2) | 4.2% | 95% | 64.6 (49.8, 82.4) | 126.0 (105.0, 150.0) | 31.8% | 203.1 (176.1, 233.0) | 396.1 (358.0, 437.1) |

Abbreviations: CAP=community-acquired pneumonia; ARI= acute respiratory infection; RSV=respiratory syncytial virus; NP/nasal swab=nasopharyngeal (NP)/nasal swab1. Obtained from the active, population-based surveillance study. CAP incidence estimates are taking into account Jena patients only; however, all three sites contributed to the RSV positive proportion
2. Percentage increase with additional specimen type results beyond NP/nasal swab, obtained from the multi-specimen study\*
3. Assumes the RSV positive proportion is the same among CAP patients tested and untested.
4. Adjusted for the underascertainment of RSV detection using the percentage increase with additional specimen type results beyond NP/nasal swab, obtained from the multi-specimen study\*
5. Proportions of CAP among RSV-related ARI hospitalizations were obtained from the multi-specimen study\*
6. Calculated by dividing RSV-related CAP incidence by the proportion of CAP in RSV-related ARI hospitalizations
\* Aliabadi N, et al Detection of Respiratory Syncytial Virus (RSV) using nasopharyngeal specimens (NPS) alone significantly underestimates disease incidence in adults hospitalized with acute respiratory infections (ARI): an updated, pooled analysis from North America. 8th Global ReSViNET Conference. Mumbai, India2024.

### Incidence of RSV-related CAP and ARI hospitalization

The unadjusted RSV-related CAP hospitalization rates were 1.9 (95% CI 0.2, 7.1) per 100,000 population in adults aged 18-59 years and 54.3 (95% CI 40.8, 70.8) per 100,000 population in adults aged ≥60 years, respectively. Using the age-group specific percent increase in detection associated with testing with additional specimen types (i.e., saliva, paired serology, and sputum) as compared to NP/nasal swab alone from the multispecimen study [12] (Supplemental Table S2), the adjusted incidence rate of RSV-related CAP hospitalizations was 4.7 (95% CI 1.5, 11.2) per 100,000 population for adults aged 18–59 years and 109.1 (95% CI 89.6, 131.6) per 100,000 population for those aged ≥60 years. After applying the proportions of CAP among the RSV-related ARI hospitalizations across different age groups [12], the estimated incidence rates of RSV-related ARI hospitalizations (adjusted) were 18.4 (95% CI 11.0, 28.9) per 100,000 population for adults aged 18-59 years, and 377.6 (95% CI 340.5, 417.7) per 100,000 population for adults aged ≥60 years (Table 2). Incidence estimates in Year 2 were about 3 times higher than in Year 1.

### RSV-related CAP hospitalization outcomes

Among RSV-positive CAP hospitalizations, 12.1% of patients aged ≥65 years died within 30 days of hospitalization; no deaths were observed for those aged 18–64 years among the 9 RSV-related CAP hospitalizations. Cardiovascular events (i.e., heart failure, arrhythmia, acute coronary syndrome, cerebrovascular accident, deep venous thrombosis or pulmonary embolism, and cardiovascular-related death) occurred in 11.1% and 36.4% of RSV-positive patients 18–64 and ≥65 years of age within 30 days of CAP hospitalization.

## Discussion

This analysis estimated the incidence rate of RSV-related ARI hospitalizations among adults between 2021 to 2023 using the RSV incidence derived from an active surveillance CAP study in Thuringia, Germany. The RSV positive proportion from NP/nasal swabs alone was 3.7% and increased to 7.8% when adjusted for undiagnosed infections due to use of a single NP/nasal swab compared to sampling with multiple diagnostic specimens (i.e., saliva, sputum, paired acute/convalescent serology, and NP/nasal swab). With further adjustment for RSV-related clinical presentations beyond CAP (e.g., non-pneumonic LRTI as well as chronic obstructive pulmonary disease [COPD] and congestive heart failure [CHF] exacerbations), the adjusted incidence rates of RSV-related ARI hospitalization for persons aged 18–59 years and ≥60 years were 18 and 378 per 100,000 population, respectively. Finally, RSV-related CAP was associated with a high risk for cardiovascular events within the month following CAP onset.

Results of our study are closely comparable to other RSV-related ARI hospitalization incidence rates in the literature. First, these values were similar to a recent time-series model-based study in Germany [21] (19–30, 33–51 and 236–363 cases per 100,000 person-years in adults aged 18-44, 45-59 and ≥60 years, respectively). Our estimates also aligned with incidence rates reported in two systematic literature reviews with meta-analyses that included adjustment for undiagnosed infections due to sole use of NP/nasal swab testing: one global meta-analysis of prospective studies in high-income countries [8] (70 and 347 cases per 100,000 population in adults aged 18-64 years and ≥65 years, respectively), and a US meta-analysis (19, 100, and 282 cases per 100,000 population in adults aged 18-49, 50–64, and ≥65 years, respectively) [18].

We adjusted for undiagnosed RSV infections due to RSV detection relying on single NP/nasal swab testing. Most RSV disease incidence studies in adults solely use RT-PCR of a NP/nasal swab for RSV detection [8, 18], but multiple studies document that single specimen testing misses RSV infections, and that adding an additional specimen such as sputum and paired serology increases detection approximately 50% [16]. The multispecimen study documented that adding multiple rather than a single specimen to NP/nasal swab approximately doubled RSV detection [12, 15]. Adjustment of adult RSV disease incidence estimates for diagnostic testing sensitivity has been incorporated in multiple recent studies, including meta-analyses of RSV disease rates in high-income countries [8] and the United States [18] as well as in recent US CDC incidence analyses [26]. Notably, the latter two studies only accounted for the percentage increase in detection rate with adding one more specimen (1.3 to 1.5-fold) whereas the most recent global study, similar to ours, included adjustment factors of ∼2 to account for the additional infections that would be identified through the synergistic effect of adding multiple additional specimens (i.e., saliva, sputum, and paired serology).

In our study, more than one third of patients aged 65 years or older with RSV-related CAP developed a cardiovascular event. Our results confirm previous observations of a high risk for cardiovascular events following RSV infections in older adults consistent with more extensively studied respiratory infections such as influenza and SARS-CoV-2 [27-29]. Time-series modelling has also documented a linkage between RSV and acute cardiac events [30]. A recent time-series study of hospitalization incidence in Germany found that 2–3% of cardiovascular hospitalizations could be attributed to RSV infection [21].

Our study had several strengths. Broad screening criteria allowed for near complete case capture and collection of minimal data on non-enrolled patients with study-qualifying CAP allowed for accurate incidence estimations. Among enrolled patients, testing rates were high (93%). A further strength of our study is the precise definition and consistent use of the catchment area population based on residential zip codes, which allowed for accurate estimations of RSV-related CAP incidence among adults.

The study also has several limitations. First, the estimation of RSV-related ARI hospitalization incidence relies on the proportions of CAP among RSV-related ARI hospitalizations among adults aged ≥40 years in the multispecimen study conducted in North America and may not be fully applicable to the German population [12]. Available literature suggests that the estimate that CAP is 28% of RSV-related ARI hospitalization [12] is conservative, as other publications have reported percentages as low as 14% [4], which would have resulted in higher incidence estimates. Second, diagnostic testing sensitivity adjustment in the current study was based on results from a North American population 40 years and older. However, a recent systematic literature found multiple studies documenting the reduced sensitivity of single NP/nasal swab PCR testing and similar results were seen among different adult age groups and geographical settings [16]. Further, minimal differences by age in the underascertainment of single specimen were seen within the study itself and this is the largest study of its kind with 3,669 participants tested and 2 countries represented (US and Canada) [12]. Third, we assumed that CAP participants who received RSV testing would have the same RSV detection rate as those who were not tested. Untested persons, especially those that could not be enrolled in the study because of lack of informed consent may have had a different detection rate since they tended to be slightly older but were less likely to be at high-risk. However, the RSV detection rates were observed to be similar across risk status (3.3% for low-risk, 3.5% for at-risk and 3.9% for high-risk). Fourth, incidence rates were calculated for Jena only and the city’s catchment population is somewhat younger than the national average, potentially leading to lower RSV incidence than would have been see in Germany as a whole as RSV-related hospitalization incidence is known to increase with age [18,19]. Lastly, the surveillance period in the CAP incidence study coincided with the COVID-19 pandemic and its immediate aftermath with larger fluctuations of incidence, which may not reflect future RSV incidence.

While RSV burden estimates relying on standard-of care testing or hospital discharge codes result in a large underestimation of the RSV-related ARI incidence, our study based on prospective, active population-based surveillance and adjustment for underascertainment provides more accurate and complete estimates. The high risk of case-fatality and cardiovascular events after RSV-related CAP confirms similar observations from other studies with larger sample size. The study results underscore the substantial burden of RSV among adults in Germany. This evidence is essential for the evaluation and implementation of novel antiviral agents for treatment and vaccines for prevention of infections due to RSV.

## Acknowledgements

Steffi Kolanos led the study nurse team and were responsible for the local logistics of the study. Susana Cubillos de Schmeer took over the management of the study team in the final phase of the study. The study nurses Stephanie Beier, Jana Schmidt, Janine Wittig and Claudia Merbold enrolled the patients.

## Data availability

All data generated or analyzed during this study are included in this published article and supplementary materials.

## Ethical statement

All study participants or legal representatives were informed about the study and consented to the participation. The study was approved by the Ethics Committee of the Jena University Hospital on 19 November 2020 (reg.no. “2020-1947-material”) and also by the Landesärztekammer Thüringen (local ethics committee) on 22 December 2020.

## Authors’ contributions

C.L., E.B., B.D.G., C.T., and M.P. contributed to conception and/or study design. C.L., E.B., S.H., L.W., Q. L., C.T., and M.P. were involved in acquisition of data, analysis and interpretation of data. C.L., E.B., S.H., J.A., L.W., C.S., L.J.B., C.V.E., Q.L., J.S., J.V., S.U., B.D.G., C.T., and M.P. participated in drafting or revision of the submitted article.

