## Supplemental Materials for "Estimated Incidence of Respiratory Syncytial Virus (RSV)-related Hospitalizations for Acute Respiratory Infections (ARIs), including Community Acquired Pneumonia (CAP), in Adults in Germany"

**Supplemental Table S1. Characteristics of adults with CAP hospitalized by RSV status [1]**

| Characteristics | RSV Positive | | RSV Negative | | | RSV Positive Proportion, |
|---|---|---|---|---|---|---|
| | N | % | N | % | P value | % |
| **Overall** | 38 | 100 | 1002 | 100 | | 3.7 |
| **Age group (Years)** | | | | | | |
| 18-59 | 6 | 15.8 | 246 | 24.6 | 0.216 | 2.4 |
| ≥60 | 32 | 84.2 | 756 | 75.4 | 0.216 | 4.1 |
| 18-64 | 9 | 23.7 | 347 | 34.6 | 0.163 | 2.5 |
| ≥65 | 29 | 76.3 | 655 | 65.4 | 0.163 | 4.2 |
| **Sex** | | | | | | |
| Female | 26 | 68.4 | 399 | 39.8 | 0.000 | 6.1 |
| Male | 12 | 31.6 | 603 | 60.2 | | 2.0 |
| **Study period [2]** | | | | | | |
| Year 1 | 14 | 36.8 | 590 | 58.9 | 0.007 | 2.3 |
| Year 2 | 24 | 63.2 | 412 | 41.1 | 0.007 | 5.5 |
| Overall study period | 38 | 100 | 1002 | 100 | | 3.7 |
| **Risk status [3]** | | | | | | |
| **Low-risk** | 3 | 7.9 | 89 | 8.9 | 1.000 | 3.3 |
| **At-risk** | 16 | 42.1 | 445 | 44.4 | 0.779 | 3.5 |
| Asthma | 1 | 2.6 | 51 | 5.1 | 1.000 | 1.9 |
| Chronic obstructive pulmonary disease | 2 | 5.3 | 80 | 8 | 0.762 | 2.4 |
| Other chronic lung disease | 1 | 2.6 | 53 | 5.3 | 0.717 | 1.9 |
| Chronic heart failure | 2 | 5.3 | 69 | 6.9 | 1.000 | 2.8 |
| Other chronic heart disease | 10 | 26.3 | 172 | 17.2 | 0.145 | 5.5 |
| Coronary artery disease | 5 | 13.2 | 74 | 7.4 | 0.202 | 6.3 |
| Chronic liver disease | 1 | 2.6 | 28 | 2.8 | 1.000 | 3.4 |
| Diabetes mellitus | 5 | 13.2 | 123 | 12.3 | 0.803 | 3.9 |
| Neurologic disease | 5 | 13.2 | 121 | 12.1 | 0.800 | 4.0 |
| Smoking | 0 | 0 | 77 | 7.7 | 0.106 | 0.0 |
| Alcoholism | 3 | 7.9 | 140 | 14 | 0.468 | 2.1 |
| **High-risk** | 19 | 50 | 468 | 46.7 | 0.690 | 3.9 |
| Asplenia | 0 | 0 | 3 | 0.3 | 1.000 | 0.0 |
| Solid malignancy | 12 | 31.6 | 247 | 24.7 | 0.332 | 4.6 |
| Hematologic malignancy | 2 | 5.3 | 77 | 7.7 | 1.000 | 2.5 |
| Chronic kidney disease | 10 | 26.3 | 226 | 22.6 | 0.587 | 4.2 |
| Immunosuppressant therapy | 11 | 28.9 | 181 | 18.1 | 0.090 | 5.7 |
| HIV/AIDS | 0 | 0 | 3 | 0.3 | 1.000 | 0.0 |
| Organ transplantation | 1 | 2.6 | 46 | 4.6 | 1 | 2.1 |
| Other Immunodeficiency | 1 | 2.6 | 22 | 2.2 | 0.579 | 4.3 |

1. Samples collected include NP/nasal swabs.
2. Year 1: 01Jul2021 – 30Jun2022; Year 2: 01Jul2022 – 30Jun2023
3. Risk factors were selected based on the overlap between the RSV and CAP studies. Low-risk was defined as patients with neither of the conditions listed under at-risk nor for high-risk at baseline

**Supplemental Table S2. Percent increase in RSV detection associated with using additional specimen type results (saliva, sputum, and paired serology) beyond NP/nasal swab [1].**

| Age group (years) | Participants with NP/nasal swab and ≥1 additional specimen | NP swab/nasal positive, n | Any specimen positive, n | Percentage increase, % | Detection rate ratio | 95% CI, % |
|---|---|---|---|---|---|---|
| ≥40 (overall) | 3669 | 120 | 254 | 112% | 2.12 | 1.86–2.41 |
| 40–59 | 1048 | 27 | 67 | 148% | 2.48 | 1.85–3.32 |
| ≥60 | 2621 | 93 | 187 | 101% | 2.01 | 1.74–2.32 |
| 40–64 | 1529 | 44 | 106 | 141% | 2.41 | 1.92–3.02 |
| ≥65 | 2140 | 76 | 148 | 95% | 1.95 | 1.66–2.28 |

1. Data from Aliabadi N, Ramirez J, McGeer A, Liu Q, Carrico R, Mubareka S, et al., editors. Detection of Respiratory Syncytial Virus (RSV) using nasopharyngeal specimens (NPS) alone significantly underestimates disease incidence in adults hospitalized with acute respiratory infections (ARI): an updated, pooled analysis from North America. Poster presentation. 8th Global ReSViNET Conference on Novel RSV Preventive and Therapeutic Interventions; 2024 February 13-16, 2024 Mumbai, India.
